# Characteristics of scientific articles on COVID-19 published during the initial three months of the pandemic: a meta-epidemiological study

**DOI:** 10.1101/2020.04.20.20073130

**Authors:** Nicola Di Girolamo, Reint Meursinge Reynders

## Abstract

**Importance:** The COVID-19 pandemic has been characterized by an unprecedented amount of published scientific articles.

**Objective:** To assess the characteristics of articles published during the first 3 months of the COVID-19 pandemic and to compare it with articles published during 2009 H1N1 swine influenza pandemic.

**Data sources:** Articles on COVID-19 and on H1N1 swine influenza indexed in PubMed (Medline) during the first 3 months of these pandemics.

**Study selection:** Any article published in the respective study periods that included any terminology related to COVID-19 or H1N1 in the title, abstract or full-text was eligible for inclusion. Articles that did not present an English abstract, as well as correspondence to previous research and erratum were excluded.

**Data Extraction and Synthesis:** Two operators conducted the selection of articles and data extraction procedures independently. The article is reported following STROBE guidelines for observational studies.

**Main Outcomes and Measures:** Prevalence of primary and secondary articles. Prevalence of reporting of limitations in the abstracts.

**Results:** Of the 2482 articles retrieved, 1165 were included. Approximately half of them were secondary articles (575, 49.4%). Common primary articles were: human medical research (340, 59.1%), in silico studies (182, 31.7%) and in vitro studies (26, 4.5%). Of the human medical research, the vast majority were observational studies and cases series, followed by single case reports and one randomized controlled trial. Secondary articles were mainly reviews, viewpoints and editorials (373, 63.2%). The second largest category was guidelines or guidance articles, including 193 articles (32.7%), of which 169 were indications for specific departments, patients or procedures. Limitations were reported in 42 out of 1165 abstracts (3.6%), with 10 abstracts reporting actual methodological limitations.

In a similar timeframe in 2009 there were 223 articles published on the H1N1 pandemic. As compared to that pandemic, during COVID-19 there were higher chances to publish reviews and guidance articles and lower chances to publish in vitro and animal research studies.

**Conclusions and Relevance:** As compared to the most recent pandemic, there is an overwhelming amount of information published on COVID-19. However, the majority of the articles published do not add significant information, possibly diluting the original information published. Also, only a negligible number of published articles reports limitations in the abstracts, hindering a rapid interpretation of their shortcomings.

**Protocol Registration:** Our protocol was registered in Open Science Framework: https://osf.io/eanzr

**KEY POINTS:** *Question:* Patients, health care professionals, policy makers, and the general public want to know what has been published on COVID-19 and what quality of research was available for decision making.

*Findings:* Half of the publications with an abstract were original research studies, i.e., for every original research article (primary article) on COVID-19 there was at least one other article that discussed or summarized what was already known (secondary article). Only 3.6% of the abstracts reported a clear statement on the limitations of the article.

*Meaning:* Clinicians and policy makers have to filter out a large body of secondary articles, which may slow down decision making during the COVID-19 pandemic.

## INTRODUCTION

Past coronavirus outbreaks have led to prolific publishing on these health issues.^1^ Similar surges in publication numbers were seen with earlier outbreaks of viral diseases like SARS, MERS, Ebola, and Swine Flu, which then dropped drastically when these diseases were contained.^1^ The production of a large bulk of literature in the early phases of such outbreaks can create a severe burden for policy makers who need to make rapid evidence-based decisions for controlling the pandemic. They have to scrutinize large quantities of scientific publications to assess what original research has been published on this topic and appraise the quality of this research. It is especially important to identify articles that report novel information to articles that summarize or comment on existing information, ie primary vs secondary articles.

In this research study we have replicated this process and report on the characteristics of articles published in the first trimester of the COVID-19 pandemic. Patients, health care professionals, policy makers, and the general public want to know what has been published on this health issue and what quality of research was available for decision making. Researchers, editors, peer reviewers, and publishing companies get an insight into the quantity and quality of articles to who’s publication they contributed. The purpose of the present meta-epidemiological study is to identify the proportion of primary and secondary articles, to identify the proportion of studies that report limitations in their abstracts and to compare publishing patterns during COVID-19 and during the previous pandemic of the XI century, the 2009 H1N1 swine influenza.

## METHODS

We performed a cross-sectional study of articles published during the initial period of the COVID-19 pandemic. We adopted the Strengthening the Reporting of Observational Studies in Epidemiology (STROBE) Statement^3^ for reporting this study and included its checklist (**Additional file 1**). We implemented 1 change compared with our original protocol. We did not assess whether studies originated as multi center research projects, because this information could not be extracted reliably.

### Eligibility criteria & Search strategy

All articles retrieved on Medline through searching PubMed with the string “(COVID-19 OR COVID)” on April 2nd 2020 at 900pm Central Standard Time, after application of the filter ‘Abstract’, were eligible for inclusion in the study. The full search strategy is given in **Additional file 2A**. Any type of article published on COVID-19 was eligible. This implies that a broad spectrum of articles ranging from letters to the editors to randomized controlled trials were eligible for inclusion. Articles were eligible if they included any terminology related to SARS-CoV-2 (including but not limited to: SARS-CoV-2, COVID, COVID-19, novel coronavirus 2019), in the title, abstract or full-text. For example, in vitro articles in which other viruses (eg, MERS-CoV, SARS-CoV) were used as a proxy for SARS-CoV-2, were still eligible for inclusion in the study as soon as the authors mentioned SARS-CoV-2 or synonyms in the manuscript. No eligibility criteria were applied to specific participants, interventions, comparators, outcomes, endpoints, language or settings of the articles. Articles that did not present an English abstract, as well as correspondence to previous research and erratum were excluded.

### Selection of articles and data extraction

We extracted from each article the following information: ‘title’, ‘abstract’, ‘DOI’’, ‘number of authors’, ‘journal’, ‘date of creation’, ‘first author’, ‘country of the first institution of the first author’, ‘article type’ (primary/secondary, defined below), ‘study design’ (defined below), ‘number of patients included’ (only for human medical research), ‘presence of objective in the abstract’, ‘presence of limitation in the abstract’, ‘main conclusion’ (**Additional file 2B**). Two operators (ND and RMR) conducted the selection of articles and data extraction procedures independently. These procedures were pilot tested on 40 articles to calibrate both operators and to fine-tune the data extraction forms. Disagreements during the selection of articles and data extraction procedures were resolved through discussions between both operators. Consultation with a third operator in the case of persisting disagreements was not necessary.

### Classifications of the included articles

We used a multi-step approach in order to classify each article included in the study. The overarching final classification was whether an article was primary, i.e., adding original scientific information to the literature, or secondary. Primary articles refer to original research studies and secondary articles refer to perspectives and syntheses of the available knowledge on COVID-19 such as, viewpoints, commentaries, guidelines, reviews etc. (**Table 1**). Our classification of included articles was not exclusively based on the labels assigned to these articles, because study designs are often mislabeled by the authors themselves.^4^ We therefore first assessed the validity of such labeling by evaluating the study design in the full-text, before making our final classifications of a study. Primary articles were divided in five categories, i.e., human medical research, in silico, in vitro, animal research and human non-medical research, and then in subcategories (**Table 1**). Many published articles included multiple analytical steps and could fall in several of these categories. For example, in a study samples could be obtained from several patients – human medical research – then transferred to a petri dish and cultured – in vitro research – then the results of the growth are modelled using computer simulation – in silico research. Since the purpose of the present meta-epidemiological study is to define the amount of information accrued that is actually relevant for healthcare policy makers and clinicians, the categorization of the articles was performed considering the theoretical order of evidence provided by different study settings, i.e., human medical research>animal research>in vitro research>in silico research. Therefore, if a study could fit in multiple categories, we assigned the highest category based on that order. In the example above, it would have been categorized as ‘human medical research’. Similarly, a study including abundant in vitro (or in silico) research and a final part on an animal model would have been categorized as ‘animal research’.

**Table 1.** Criteria employed to classify the included articles.

| Article type | Study design | Description |
| --- | --- | --- |
| Primary articles | Human medical research | Human medical research refers to articles reporting information on 1 or more human patient/s. In order to be classified as human medical research, an article would need to report individual patient data. Articles in this category were further subcategorized in ‘randomized controlled trials (RCTs)’, ‘observational studies and case series’, and ‘case reports’, based on the following key: articles including a single case were categorized ‘case reports’; articles including 2 or more cases where no randomization were performed were categorized ‘observational studies and case series’; articles including 2 or more cases where randomization of a treatment was performed were categorized ‘RCTs’. We extracted the total number of patients included in human medical research studies. |
|  | In silico research | Primary articles were classified as ‘in silico research’ if they reported the results of any type of computer-based research. Articles in this category were further subcategorized in ‘epidemiological modelling’, ‘biology/biochemistry/bioinformatics studies’, and ‘social media studies’, based on the following key: articles focusing on exploiting platforms or other online tools, such as Google trend in order to extrapolate information or generate any sort of prediction were categorized ‘social media studies’; articles using published or original data to calculate the spreading or |
|  |  | impact of COVID-19, including but not limited to epidemiological models and calculations, were classified as ‘epidemiological modelling’; articles using published or original data in order to generate original information solely using computer processing in the field of biology, biochemistry and bioinformatics were classified as ‘biology/biochemistry/bioinformatics studies’. If any part of the work performed by researchers was done without the computer the studies would have been included in the categories in vitro, human medical research or animal research. |
|  | In vitro research | Primary articles were classified as ‘in vitro research’ if they reported the results of any type of laboratory-based or in vitro research without inclusion of human or animal subjects. Articles in this category were further subcategorized in ‘development/performance of diagnostic technology’, ‘virus-host interaction’, ‘genomic studies’, ‘pharmacological activity in vitro’, ‘viral isolation/transport/elimination’ based on their primary objective and results. |
|  | Animal research | Research including animal subject/s refers to original clinical research on 1 or more animal subject/s. This category was not further subdivided and individual results of the included studies were reported. |
|  | Human non-medical research | Primary articles were classified as ‘human non-medical research’ if they reported the result of surveys or studies performed on |
|  |  | healthcare professionals. Articles classified as a ‘survey’ were subdivided in ‘surveys to health professionals’ and ‘surveys to the general public’ depending on the population surveyed. |
| Secondary articles | Systematic review | Secondary articles were classified as ‘systematic review’ if they reported the results of a systematic search, whether a narrative review or a meta-analysis was present. |
|  | Review/viewpoint/editorial/letter/news | Secondary articles were classified as ‘review/viewpoint/editorial/letter/news’ if they reported a narrative or graphical representation of previously performed research or previously published information. Since we found it difficult to unequivocally distinguish between articles in this category, the articles were not further subdivided. |
|  | Guideline/guidance/recommendation | Secondary articles were classified as ‘guideline/guidance/recommendation’ if they reported guidelines or recommendations either on the basis of personal experience, a review of the literature, or a combination of them. This category was further classified in ‘indications for specific department/disease/procedure’ if they reported clinical recommendations for a subset of health professionals and ‘indications for lay public’ if they reported recommendations/indications for the general public. |
|  | Correspondence to previous research | Secondary articles were classified as ‘correspondence to previous research’ if they reported any type of direct commentary to a |
|  |  | recently published article. |
|  | Erratum/correction | Secondary articles were classified as 'erratum/correction' if they reported a mistake with or without a correction for a previously published article. |

### Abstract assessment

We screened all abstracts to assess whether the objectives and the limitations of the article were reported or not. An abstract was defined as any type of information reported in the area for abstracts in PubMed. Objectives were defined as ‘reported’ when the abstract reported any type of statement that explained the purpose of the article. Limitations were defined as ‘reported’ when the abstract reported any type of statement that explained limitation(s) of the article. Limitations were further subdivided in ‘methodological limitation’ and ‘general limitation’; articles were classified as reporting a ‘methodological limitation’ when they stated in the abstract the presence of at least 1 limitation inherent to the article design (e.g., “due to the inclusion of a convenient sample this report is at risk of selection bias”); articles were classified as reporting a ‘general limitation’ when they stated in the abstract the presence of a limitation that was not inherent to the article’s design (e.g., “more evidence is needed”, “further research on the topic is warranted”).

### Selection, extraction & classification of articles on H1N1 2009 pandemic

We performed a search & extraction in an analogous way for articles published during the early phases of the H1N1 2009 pandemic. We performed a search on Medline through PubMed with the string “H1N1”. We applied the filter regarding Text Availability “Abstract” and ordered the articles by date of publication. Our search strategy is reported in Additional file 2C. We extracted all the articles retrieved through the “Save” function on a .csv file. We established which was the first published article on the H1N1 2009 pandemic^5^ based on a CDC summary.^6^ We then included three full months of publications, ie from April 25^th^ 2009 to July 25^th^ 2009. Similar to articles related to COVID-19, articles were eligible for inclusion if they reported terminology related to “H1N1”, “swine flu” or “the current pandemic”, among others. From the articles included, we extracted country of origin, language of full-text, type of study and study design were extracted in a similar fashion as was done for the COVID-19 articles. The selection, extraction and classification of articles was performed independently by two operators (ND and RMR) and disagreements were resolved by consensus.

### Outcomes and prioritization

The primary outcomes of this meta-epidemiological study were:

- The proportion of primary articles over the total number of articles with an abstract published during the first three months of the COVID-19 pandemic.
- The proportion of articles reporting limitations in their abstracts.
- The proportion of article types during COVID-19 and during the 2009 H1N1 swine flu pandemic.

The associations of any of these outcomes with other individual article characteristics were secondary outcomes.

### Statistical analysis

Descriptive statistics are expressed as medians with interquartile ranges (IQR) and ranges or absolute counts and percentages. Multivariable logistic regression models were developed to explore the factors associated with the primary outcomes and provide odds ratio adjusted for confounders. Variables retained clinically significant were entered in the models regardless of their statistical significance. Goodness of fit was assessed with Hosmer-Lemeshow test and Nagelkerke R squared was used as a measure of predictive power. The first multivariable logistic regression had primary vs secondary articles as the dependent variable and included the country of publication (limited to the 11 countries with more publications), the language of full-text (English/Other languages), the number of authors, and the number of days from the start of the pandemic as predictor variables. The initial model had a significant Hosmer-Lemeshow test (P=0.004) and a low Nagelkerke R squared (0.27), due to non-linearities in the number of authors variable. The model was rebuilt after binning the variable (0 authors, 1-2 authors, 3-5 authors, 6-10 authors, >11 authors). The new model had a non-significant Hosmer-Lemeshow test (P=0.14) and higher Nagelkerke R squared (0.33) and was retained. A univariable logistic regression model was built including COVID-19 articles vs H1N1 articles as the dependent variable and including article type as predictor variable.

Data analyses and figures were performed using SPSS (version 24, IBM) and R 3.6.3 (R Core Team, 2020, www.R-project.org/). All P values were two tailed with nominal statistical significance claimed for P <0.05.

## RESULTS

### Results of the search

The results of our search are presented in a flow diagram (**Figure 1**). Our search yielded 2482 articles. After exclusions of articles without an abstract, we retrieved 1215 articles. We excluded 50 articles based on the following rationale: duplicate articles (n=13), articles that were not on COVID-19 (13), articles without an English abstract (6), letter to previous papers (8), erratum (2), local morbidity reports (7), and statement of the WHO (1). We included a total of 1165 articles on COVID-19 in the study.

**Figure 1.**
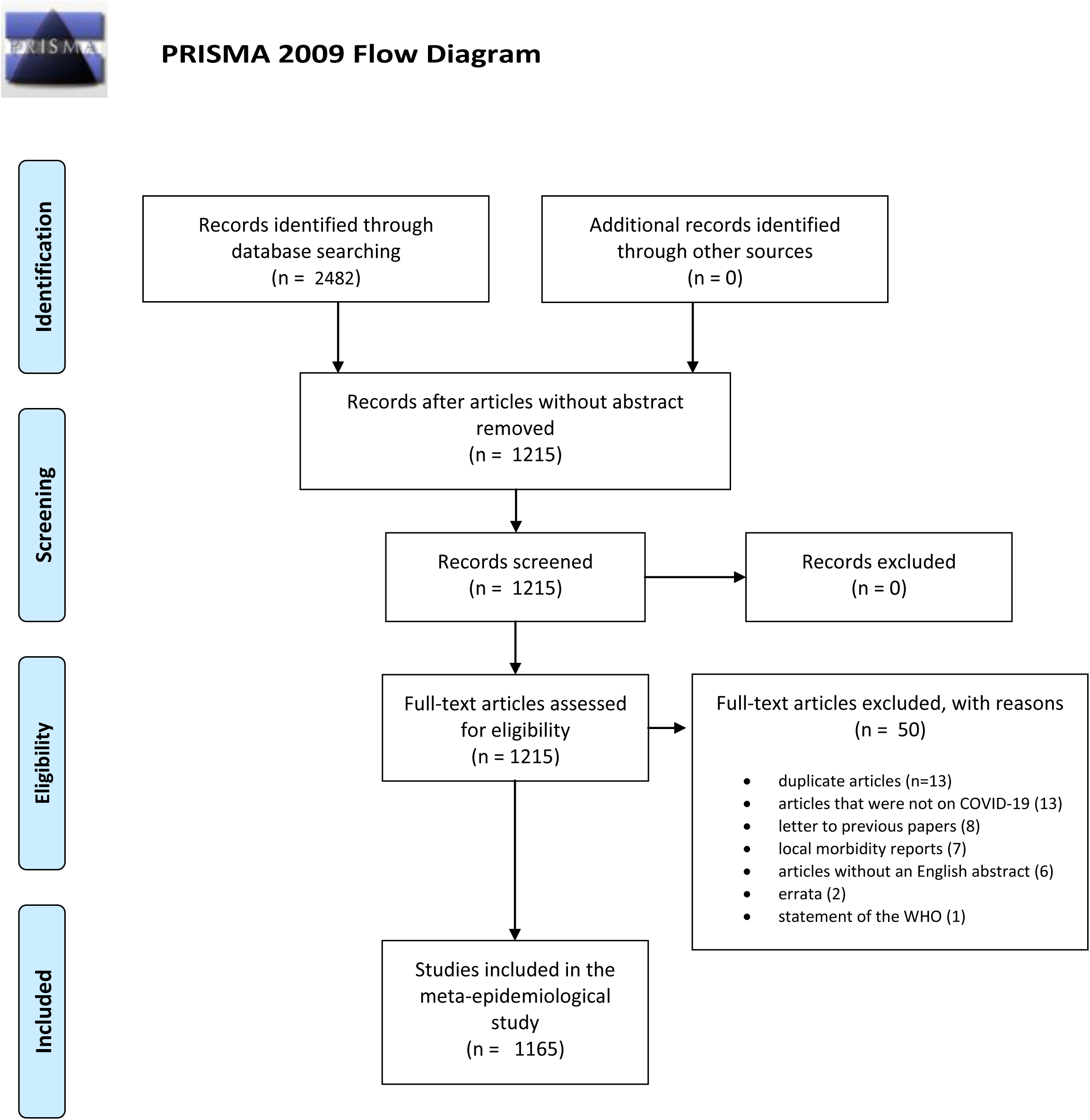
Modified PRISMA flow diagram showing the article inclusion process.

### Characteristics of published articles

Four countries contributed to three quarters (871, 74.8%) of the included articles, i.e., approximately half of these articles, (588, 50.5%) came from China, 168 articles (14.4%) from the United States, 77 articles (6.6%) from Italy, 38 articles (3.3%) from the United Kingdom. The remaining 294 articles (25.2%) originated in decreasing numbers in Japan, Singapore, Korea, India, France, Germany, Taiwan, and other countries (**Table 2**).

**Table 2.**
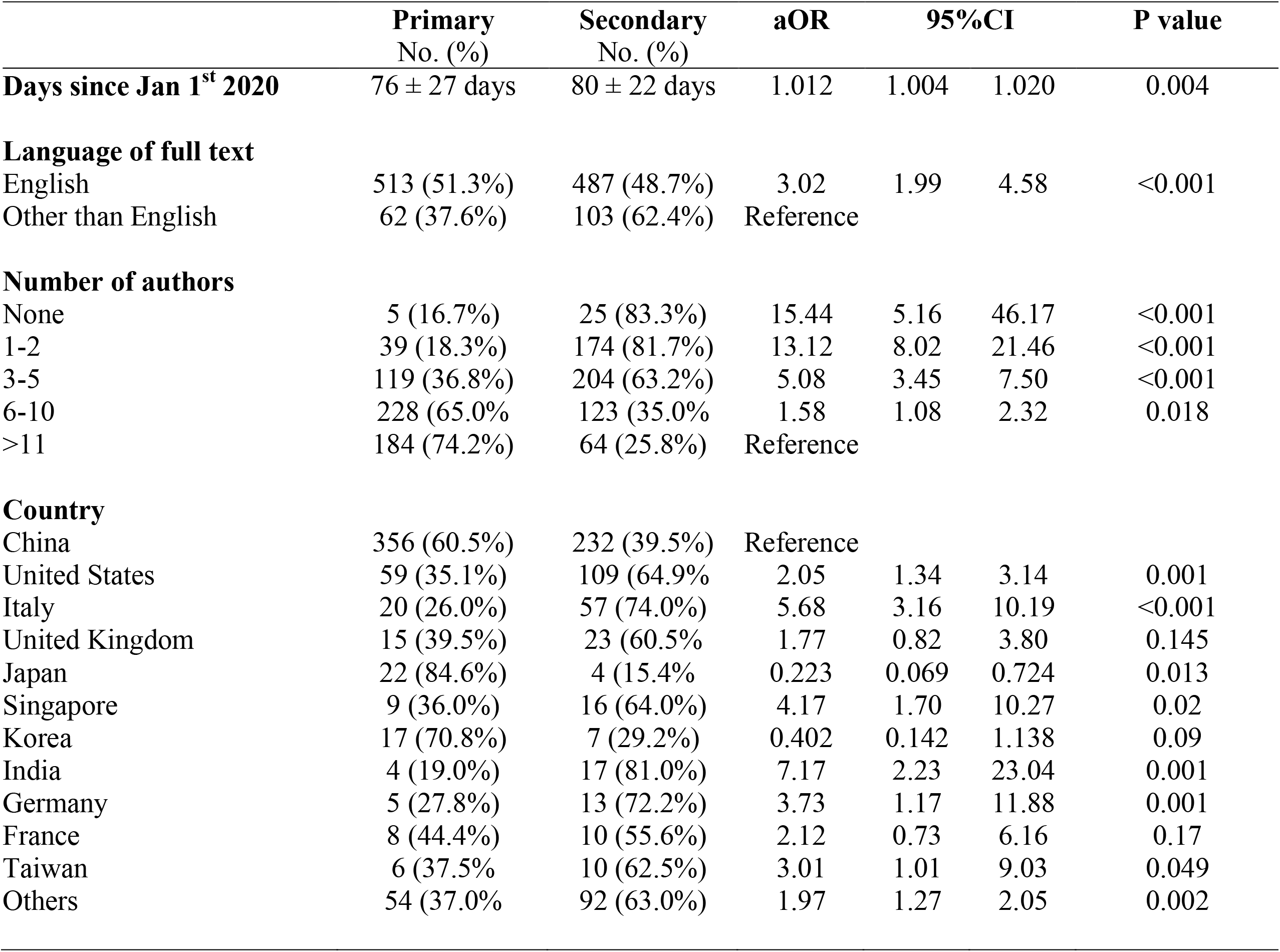
Results of multivariable logistic regression analysis to determine factors associated with primary article publication in 1165 articles published in the early stages of COVID-19 pandemic. Continuous data are reported as Median ± IQR. Binary data are reported as number of observed events (percentage over the total). Hosmer-Lemeshow test: Chi-square=12.2; P=0.14. Nagelkerke R squared: 0.33. aOR: adjusted odds ratios; CI: Confidence intervals.

|  | Primary<br>No. (%) | Secondary<br>No. (%) | aOR | 95%CI |  | P value |
| --- | --- | --- | --- | --- | --- | --- |
| **Days since Jan 1<sup>st</sup> 2020** | 76 ± 27 days | 80 ± 22 days | 1.012 | 1.004 | 1.020 | 0.004 |
| **Language of full text** |  |  |  |  |  |  |
| English | 513 (51.3%) | 487 (48.7%) | 3.02 | 1.99 | 4.58 | <0.001 |
| Other than English | 62 (37.6%) | 103 (62.4%) | Reference |  |  |  |
| **Number of authors** |  |  |  |  |  |  |
| None | 5 (16.7%) | 25 (83.3%) | 15.44 | 5.16 | 46.17 | <0.001 |
| 1-2 | 39 (18.3%) | 174 (81.7%) | 13.12 | 8.02 | 21.46 | <0.001 |
| 3-5 | 119 (36.8%) | 204 (63.2%) | 5.08 | 3.45 | 7.50 | <0.001 |
| 6-10 | 228 (65.0%) | 123 (35.0%) | 1.58 | 1.08 | 2.32 | 0.018 |
| >11 | 184 (74.2%) | 64 (25.8%) | Reference |  |  |  |
| **Country** |  |  |  |  |  |  |
| China | 356 (60.5%) | 232 (39.5%) | Reference |  |  |  |
| United States | 59 (35.1%) | 109 (64.9%) | 2.05 | 1.34 | 3.14 | 0.001 |
| Italy | 20 (26.0%) | 57 (74.0%) | 5.68 | 3.16 | 10.19 | <0.001 |
| United Kingdom | 15 (39.5%) | 23 (60.5%) | 1.77 | 0.82 | 3.80 | 0.145 |
| Japan | 22 (84.6%) | 4 (15.4%) | 0.223 | 0.069 | 0.724 | 0.013 |
| Singapore | 9 (36.0%) | 16 (64.0%) | 4.17 | 1.70 | 10.27 | 0.02 |
| Korea | 17 (70.8%) | 7 (29.2%) | 0.402 | 0.142 | 1.138 | 0.09 |
| India | 4 (19.0%) | 17 (81.0%) | 7.17 | 2.23 | 23.04 | 0.001 |
| Germany | 5 (27.8%) | 13 (72.2%) | 3.73 | 1.17 | 11.88 | 0.001 |
| France | 8 (44.4%) | 10 (55.6%) | 2.12 | 0.73 | 6.16 | 0.17 |
| Taiwan | 6 (37.5%) | 10 (62.5%) | 3.01 | 1.01 | 9.03 | 0.049 |
| Others | 54 (37.0%) | 92 (63.0%) | 1.97 | 1.27 | 2.05 | 0.002 |

Half of the included articles, (578, 49.6%) came from 49 individual journals (range of published articles per journal, 5-70). The full-text of 1000 of the 1165 articles (85.8%) was in English. Of the remaining articles, 152 full-texts (13.0%) were in Chinese, 6 (0.5%) in Spanish, 5 (0.4%) in German, and 2 (0.2%) in French. Articles included an average of 7.4 authors (SD: 6.98), ranging from 0 to 65 authors.

We identified 590 (50.6%) primary and 575 (49.4%) secondary articles. Of the primary articles, 340 were human medical research (59.1%), 182 were in silico studies (31.7%), 26 were in vitro studies (4.5%), 20 were human non-medical research (3.5%), and 7 were animal research (1.2%). Of the secondary articles, the majority were reviews, viewpoints and editorials (373, 63.2%). The second largest category was guidelines or guidance articles, including 193 articles (32.7%), of which 169 were indications for specific departments, patients or procedures. We included 23 systematic reviews (3.9%) and 1 protocol (0.2%).

Based on the multivariable logistic regression model, secondary articles were more likely to be published in a language different than English (aOR: 3.02, 95% CI: 1.99 to 4.58), to be published at a later stage of the pandemic (aOR: 1.01, 95% CI: 1.00 to 1.02), to include a lower number of authors (multiple aORs, **Table 2**), and to be published by authors from India, Italy, Singapore, Germany, and Taiwan (multiple aORs, reference: China; **Table 2**).

### Classification of primary articles

Human medical research consisted of 281 observational studies or case series (82.6%), 58 single case reports (17.1%), and 1 randomized controlled trial (0.3%). Human medical research included a median of 23 patients (IQR: 85), ranging from 1 to 72314 patients. When only observational studies and case series were considered, the median number of patients included was 38 (IQR: 106). The only RCT included in the study enrolled 199 patients.

In silico research consisted of 109 studies on epidemiological modelling (59.9%), 64 studies on biochemistry, biology, bioinformatics or molecular modelling (35.2%), 5 studies evaluating or exploiting social media (2.7%), 2 studies on economical modelling (1.6%) and 1 description of an open database for viral trends (0.5%).

In vitro research consisted of 7 studies on the development or performance of diagnostic technology (26.9%), 7 studies on virus-host interactions (26.9%), 6 studies on gene expression or genomics (23.1%), 3 studies on pharmacological activity of compounds (11.5%), and 3 studies on viral isolation, transport or elimination (11.5%).

Animal research consisted of 4 studies that included mice (1 immunization with SARS-CoV S, 1 pharmacokinetic of a α-ketoamide inhibitor, 1 viral challenge with HCoV-OC43 and treatment with EK1C4, 1 hepatectomy and consequent gene expression)(57.1%), 1 study on hamsters challenged with SARS-CoV 2 (14.3%), 1 study on macaques challenged with MERS-CoV and treated with GS-5734 (14.3%), and 1 study on presence of SARS-CoV-2 related coronaviruses in Malayan pangolins (14.3%).

Human non-medical research consisted of 15 surveys, 8 on health professionals (40%), 7 on lay public (35.0%), 2 surveys of healthcare facilities (10.0%), 1 development of a psychological scale (5.0%), 1 RCT on medical professionals (5.0%), 1 simulation of an outbreak in a hospital (5.0%).

### Reporting of limitations in the abstract

Limitations were reported in 42 out of 1165 abstracts (3.6%). Ten abstracts reported methodological limitations, i.e., limitations related to the study design and the remaining 32 abstracts reported general limitations, such as the current lack of evidence on COVID-19, or the need for further studies on COVID-19. Limitations were reported in 5 out of 23 systematic reviews (21.7%) and 2 out of 20 human non-medical researches (10.0%). All other manuscript types had a frequency of reporting limitations between 0% and 3.8%.

### Comparison with early publications during the 2009 H1N1 swine influenza pandemic

Our search yielded 434 articles. After exclusions of articles without an abstract, we retrieved 239 articles. We excluded 16 articles that did not mention H1N1 or swine influenza in the full text. We included a total of 223 articles published at early stage of the 2009 H1N1 pandemic in the study. Eight countries contributed to three quarters (166, 74.4%) of the included articles, with approximately one third of the articles coming from the United states (75, 33.6%) and one tenth of them coming from China (24, 10.8%). Almost all the articles (215, 96.4%) had an English full text. Based on our previous classification, there were 179 primary articles (80.3%) and 44 secondary articles (19.7%). The primary articles included 71 human medical researches (39.7%), 36 animal researches (20.1%), 33 in vitro studies (18.4%), 30 in silico studies (16.7%), and 9 human non-medical researches (5.0%). Of the human medical research, 66 were observational studies and case series (92.9%), 3 were RCTs (4.2%), and 2 were single case reports (2.8%). The secondary articles were mainly reviews, viewpoints and editorials (38, 86.4%), with a few guidelines or guidance articles (5, 11.4%) and 1 systematic review (2.3%)(**Table 3**).

**Table 3.** Type of studies published in the early stages of COVID-19 pandemic and of 2009 H1N1 pandemic. Results of univariable logistic regression analysis are reported. Binary data are reported as number of observed events (percentage over the total articles for each pandemic event; for human medical research subcategories, the percentage is calculated over the number of human medical researches). OR: odds ratios; CI: Confidence intervals.

|  | **COVID-19**<br>**pandemic**<br>**No. (%)** | **H1N1 2009**<br>**pandemic**<br>**No. (%)** | **OR** | **95%CI** |  | **P value** |
| --- | --- | --- | --- | --- | --- | --- |
| **Type of study** |  |  |  |  |  |  |
| Human medical research | 340 (29.2%) | 71 (31.8%) | Reference |  |  |  |
| Single case reports | 58 (17.1%) | 2 (2.8%) |  |  |  |  |
| Observational studies | 281 (82.6%) | 66 (93.0%) |  |  |  |  |
| Randomized controlled trials | 1 (0.3%) | 3 (4.2%) |  |  |  |  |
| Human non-medical research | 20 (1.7%) | 9 (4.0%) | 0.46 | 0.20 | 1.06 | 0.07 |
| In silico | 182 (15.6%) | 30 (13.5%) | 1.27 | 0.80 | 2.01 | 0.32 |
| In vitro | 26 (2.2%) | 33 (14.8%) | 0.16 | 0.09 | 0.29 | <0.001 |
| Animal research | 7 (0.6%) | 36 (16.1%) | 0.04 | 0.02 | 0.09 | <0.001 |
| Guidelines/guidance | 193 (16.6%) | 5 (2.2%) | 8.06 | 3.20 | 20.30 | <0.001 |
| Review/viewpoint/editorial/letter/news | 373 (32.0%) | 38 (17.0%) | 2.05 | 1.35 | 3.12 | 0.001 |
| Systematic review | 23 (2.0%) | 1 (0.4%) | 4.80 | 0.64 | 36.15 | 0.13 |

In the univariable logistic regression model, the odds of being published during COVID-19 were 8 times higher for guideline articles (OR: 8.1, 95% CI 3.2 to 20.3), and 2 times higher for reviews (OR: 2.0, 95% CI 1.3 to 3.1), while the odds of being published during H1N1 were 24 times higher for animal researches (OR: 24.6, 10.5 to 57.6), and 6 times higher for in vitro research (OR: 6.1, 3.4 to 10.8)(**Table 3**; **Figure 2**).

**Figure 2.**
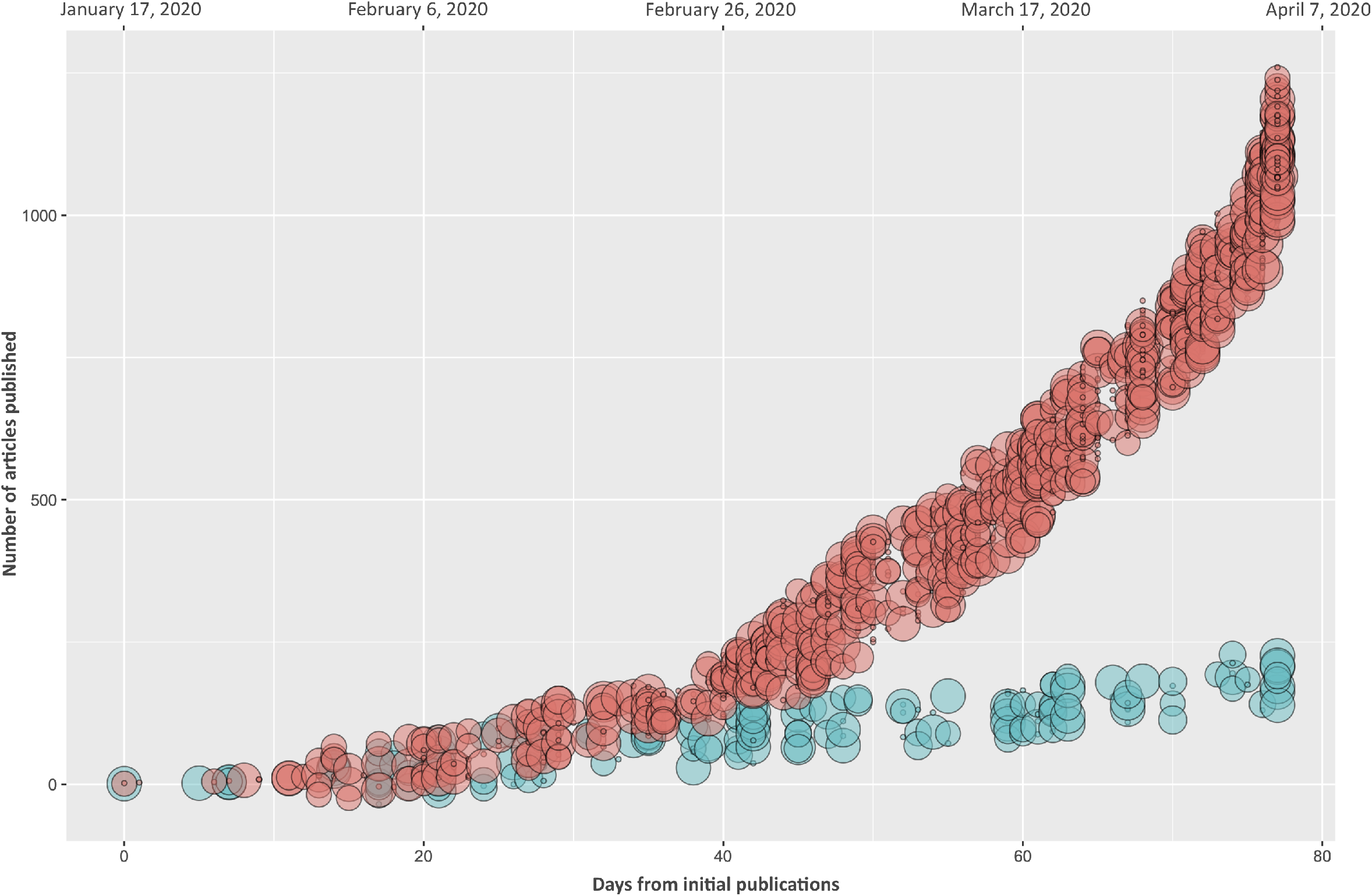
Relationship between days from the start of the pandemics, number of articles published and type of articles during COVID-19 pandemic (red circles) and 2009 H1N1 swine flu pandemic (green circles). On the × axis the days from the start of COVID-19 pandemic (top axis) and the days from the first publication for each pandemic (bottom axis) are reported. On the y axis the total number of articles published for each pandemic is reported. Circle size has been arbitrarily classified in order to show different levels of clinical evidence: at increasing circle size, increase the value of the article (circle size: 1: secondary articles and human non-medical research; 7: in silico research; 8: in vitro research; 9: animal research; 11: case reports; 14: observational studies and case series; 17: randomized controlled trials; 20: systematic reviews). The values were jittered over the y axis to reduce superimposition of data.

## DISCUSSION

### Principal findings of the study

This meta-epidemiological study is novel in having assessed the characteristics of scientific articles published during the initial three months of the COVID-19 pandemic. Our study has five key findings. First, almost half (49.4%) of all 1165 included articles were secondary articles. Perspectives and syntheses have an important role in scientific research, but one secondary article for each primary article could be redundant. Second, human medical research consisted of 29.2% (340/1165) of the included articles. This implies that a large body of articles are not relevant for health care policy makers. Identifying human medical research studies slows down the evidence-based decision making process, because a large bulk of literature has to be filtered out first. This selection process is particularly time consuming, because it can often not be done by reading titles and abstracts alone. Third, all except one (339/340) of the human medical research studies were observational studies or case reports. This implies that policy makers have to rely predominantly on studies that get a low-certainty (or quality) rating according to the GRADE (Grading of Recommendations Assessment, Development and Evaluation) approach.^7^ Fourth, about half of all included articles originated in China, i.e., 50.5% (588/1165). A high prevalence of articles from China was expected because the COVID-19 outbreak started in that country, but this statistic is disproportionate with the much higher COVID-19 infection and death rates in other countries. Fifth, only 3.6% (42/1165) of all included articles reported limitations in their abstracts. Reporting limitations is an important warning sign for end-users of research articles and is an obligatory item in the reporting of abstracts of systematic reviews.^8^

### Comparison with other studies

The exponential growth of publications as was identified in this paper during the first three months of the COVID-19 pandemic was also identified in past viral outbreaks such as SARS, MERS, Ebola, and Swine Flu.^1^ This high publication rate dropped dramatically upon containment of these diseases. Gori et al.^9^ identified a high proportion of secondary literature in the first 30 days of the COVID-19 pandemic. However, their findings cannot be directly compared with ours because they used different methods, had a much smaller sample size (234 papers versus 1165 in our sample), measured mostly different outcomes and at different time points (one month versus 3 months in our sample).

### Strengths and weaknesses

The strengths of this meta-epidemiological study are: (1) this is the first research study that assessed the characteristics of articles on COVID-19 listed in PubMed in the first three months since the outbreak of the coronavirus pandemic (2) all study selection and data extraction procedures were conducted by two methodologists independently and all raw data were reported in additional files (3) the manuscript was reported according to the STROBE checklist. Having searched eligible articles exclusively in PubMed is a limitation, because this could have biased our outcomes.^10,11^ The total body of literature on COVID is expected to be larger.

### Implications and future research

The exponential surge in scientific publishing was expected with the outbreak of a pandemic of an unknown virus. Finding mostly observational studies among the human medical research studies in this body of articles was also not surprising. However, having to filter out half of the literature, because it is not producing new research data is problematic, especially when almost 2500 new articles on COVID-19 were indexed in PubMed in the first 3 months of this pandemic. Researchers, peer reviewers, editors, and publishing companies are responsible for this large body of literature. They should aim at flattening the publication curve for example by tightening their acceptance criteria. This strategy could also help to improve the overall research quality.^12^ Labeling publications as ‘secondary article’ in the abstract could become an initial obligatory item for all publications that do not produce original research.

Undertaking future research studies on outbreaks of diseases should start with the consultation of a wide body of stakeholders to develop and prioritize research questions. Such research could explore (1) our statistics at later time points (2) quality assessments of the conduct and reporting of research studies on COVID-19 (3) factors that could be implemented to control the quantity and quality of publications (4) the impact of the development of a vaccine for COVID-19 on the publication curve and (5) how to rapidly synthesize literature in times of a pandemic. Further, high quality systematic reviews and guidelines for the prevention and management are necessary when COVID-19 is contained. This will be key to control new outbreaks of COVID-19 and other diseases.

## CONCLUSIONS

We showed that as compared to the most recent pandemic (2009 H1N1), there is an overwhelming amount of information published on COVID-19. Due to the large body of non-original research data (50%) published in the early phases of the pandemic, the original information published has been diluted. Also, a negligible number of published articles reported limitations in the abstracts, potentially facilitating overemphasis of the article findings or recommendations. Researchers, peer reviewer, and editors, should take action to level the publication curve and start labeling non-original research articles as secondary articles.

## Data Availability

Currently data are not available since more research is being performed on them.

